# COVID-19 transmission in Mainland China is associated with temperature and humidity: a time-series analysis

**DOI:** 10.1101/2020.03.30.20044099

**Authors:** Hongchao Qi, Shuang Xiao, Runye Shi, Michael P. Ward, Yue Chen, Wei Tu, Qing Su, Wenge Wang, Xinyi Wang, Zhijie Zhang

**Author notes:** Correspondence author **Correspondence:** Zhijie Zhang, Department of Epidemiology and Health Statistics, School of Public Health, Fudan University, Shanghai, China. Contributed equally (Hongchao Qi, Shuang Xiao).

## Abstract

COVID-19 has become a pandemic. The influence of meteorological factors on the transmission and spread of COVID-19 if of interest. This study sought to examine the associations of daily average temperature (AT) and relative humidity (ARH) with the daily count of COVID-19 cases in 30 Chinese provinces (in Hubei from December 1, 2019 to February 11, 2020 and in other provinces from January 20, 2020 to Februarys 11, 2020). A Generalized Additive Model (GAM) was fitted to quantify the province-specific associations between meteorological variables and the daily cases of COVID-19 during the study periods. In the model, the 14-day exponential moving averages (EMAs) of AT and ARH, and their interaction were included with time trend and health-seeking behavior adjusted. Their spatial distributions were visualized. AT and ARH showed significantly negative associations with COVID-19 with a significant interaction between them (0.04, 95% confidence interval: 0.004–0.07) in Hubei. Every 1°C increase in the AT led to a decrease in the daily confirmed cases by 36% to 57% when ARH was in the range from 67% to 85.5%. Every 1% increase in ARH led to a decrease in the daily confirmed cases by 11% to 22% when AT was in the range from 5.04°C to 8.2°C. However, these associations were not consistent throughout Mainland China.

## Introduction

Starting in December 2019, the severe acute respiratory coronavirus 2 (SARS-CoV-2) identified in Wuhan, China, has caused an outbreak of a novel coronavirus disease (COVID-19)[1]. Its typical clinical symptoms include fever, dry cough, myalgia, and pneumonia, and may cause progressive respiratory failure due to alveolar damage and death[2]. As of March 23, 2020, a total of 81,603 confirmed cases (61.28% in Wuhan) and 3,276 deaths have been reported in mainland China[3]. And now it is a pandemic [4].

Respiratory droplets and person-to-person contact are the major routes of transmission of the coronavirus[5]. Environmental factors such as temperature and relative humidity may influence the transmissions of coronavirus[6] by affecting the survival of the virus in its transmission routes, there has evidence for severe acute respiratory coronavirus (SARS-CoV)[7, 8] and Middle East respiratory syndrome coronavirus (MERS-CoV)[9, 10]. Whether and how meteorological factors affecting the spread of COVID-19 has so far not been investigated. Using data of confirmed cases from all provinces in mainland China, we investigated the correlations between meteorological factors and the daily cases of COVID-19. The goal is to provide scientific evidence regarding the future progression of COVID-19 under the changing circumstances of climate factors.

## Materials and methods

## Study area and data

Daily counts of laboratory-confirmed cases in all provinces in China were collected from the official reports of the National Health Commission of People’s Republic of China from December 1, 2019 to February 11, 2020 for Hubei province and from January 20, 2020 to February 11, 2020 for other provinces. Case definitions for suspected cases and laboratory-confirmed cases and the description of the surveillance system have been published online and reported elsewhere[11, 12]. Tibet was not included in the following model since only one case was reported during the 23-day period.

The meteorological data, including daily average temperature (AT) and daily average relative humidity (ARH) of each provincial capital, were retrieved from Weather Underground (https://www.wunderground.com/). The following keywords were applied in the Baidu index (http://index.baidu.com/), the largest search engine in China, using the keywords “Wuhan pneumonia” OR “novel coronavirus” OR “coronavirus disease 2019” OR “coronavirus disease-19” OR “2019 novel coronavirus” OR “2019-nCoV” OR “SARS-CoV-2” OR “COVID-19” OR “SARS-CoV-2” OR “severe acute respiratory coronavirus 2”. The province-based Baidu index with massive Internet behavior data recorded, including concerns for COVID-19, was used as a measure of health-seeking behavior that may affect the transmission of coronavirus[13].

### Statistical analysis

A generalized additive model (GAM) was applied to quantify the province-specific associations between meteorological variables and the daily cases of COVID-19 during the study periods, accounting for short-term temporal trend and health-seeking behavior through Baidu index.

The distribution of COVID-19 cases was assumed to be a negative binomial given that the variances of the daily counts were larger than their means. Considering the incubation period of COVID-19, the effects of AT, ARH, and covariates were modeled with a 14-day exponential moving average (EMA) to account of their potential lag effects and the interaction between AT and ARH was tested and included in the model if significant. The short-term temporal trend was indicated using natural splines of time with two degrees of freedom. The model is given by:

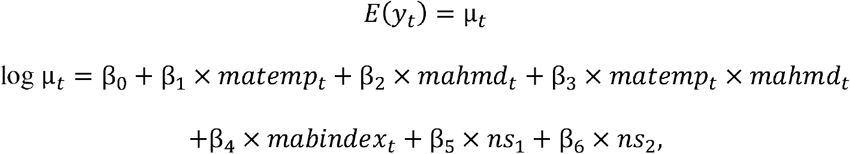

where *y*_*t*_ is the daily counts of COVID-19 at day *t*, μ_*t*_ is the expected value of daily counts at day *t*, β_0_ is the intercept, β_1_ denotes the effect of moving average of AT, β_2_ is the effect of moving average of ARH, β_3_ is the interaction between AT and ARH, β_4_ denotes the effect of moving average of Baidu index, and β_5_ and β_6_ are the regression coefficients of natural splines of time with two degrees of freedom.

Considering the effect of the interaction between AT and ARH, effect plots were shown with the change in log-transformed daily counts over AT (or ARH) given the ARH (or AT) of the 25^th^, 50^th^, and 75^th^ percentiles. Additionally, the effects of meteorological factors were parameterized with the incidence rate ratio (IRR) assuming the number of persons at risk stayed stable during the study period. IRRs of AT (or ARH) given the ARH (or AT) of the 25^th^, 50^th^, and 75^th^ percentiles in models were estimated and depicted using forest plots and geographic maps.

### Sensitivity analysis

To verify model results, a sensitivity analysis was performed in which the time-series in Wuhan and in Hubei province were restricted to the period the same as that in other provinces − i.e. January 20 to February 11 − with the same GAM fit to the data (the results of sensitivity analysis in Figs. A8-A10).

The R software (version 3.5.3, http://cran.r-project.org; R Foundation for Statistical Computing, Vienna, Austria) was used to perform statistical analyses. QGIS Desktop (version 3.4.14, https://www.qgis.org/; Open source geospatial foundation project) from the open-source geospatial foundation project was used to plot the geographic patterns.

## Results

The daily counts of confirmed cases, AT, and ARH for 31 provinces were summarized in Table A1. The cumulative confirmed cases varied from one (Tibet) to 33,453 (Hubei), and 74.7% of all confirmed cases occurred in Hubei for the periods (Fig. 1a). The average AT ranged from −16.96°C to 19.3°C (Fig. 1b), and the average ARH ranged from 17.93% to 86.20% (Fig. 1c).

**Fig 1.**
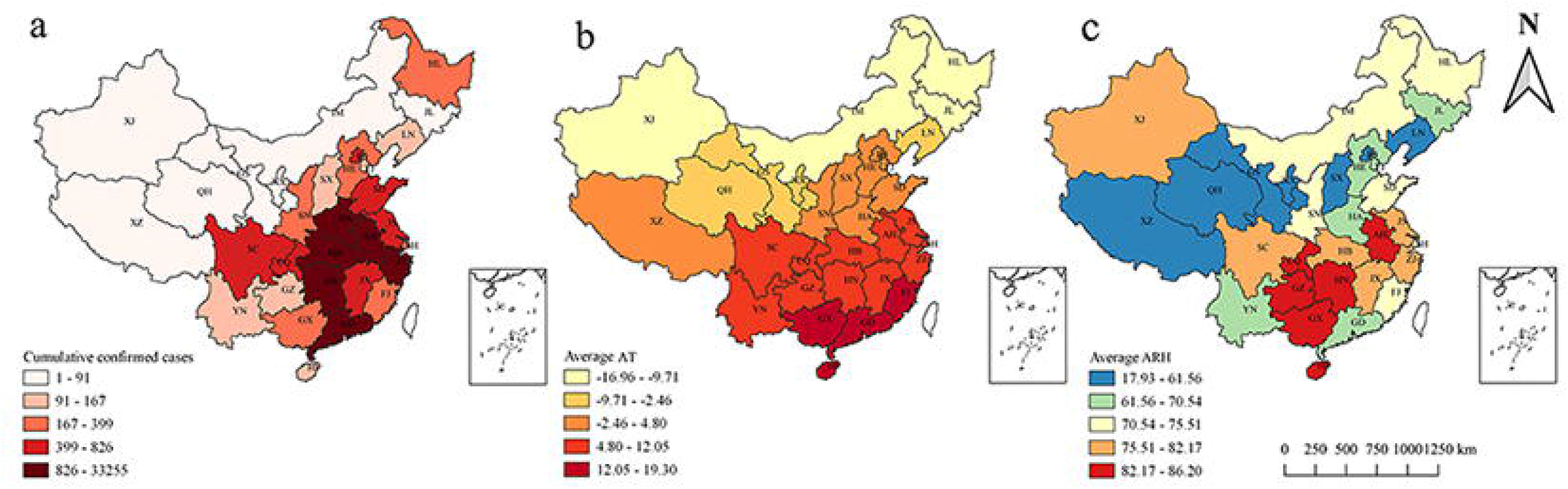
The distribution of (a) cumulative confirmed cases, (b) average AT, and (c) average ARH in all provinces surveyed in China.

Fig. 2 shows that the daily counts in Hubei increased sharply after January 20, 2020. The daily AT varied from 1.5°C to 11.42°C, fluctuating mostly around 6°C. The daily ARH varied from 42.17% to 96.92%, remaining above 70% on most days (68.49%). For other provinces, more than 88% confirmed cases occurred between January 23, 2020 and February 8, 2020, and the time series of the confirmed cases appeared in an “M” shape, especially in Guangdong and Zhejiang (Fig. A1). ARH and AT varied across provinces (Figs. A2 and A3).

**Fig 2.**
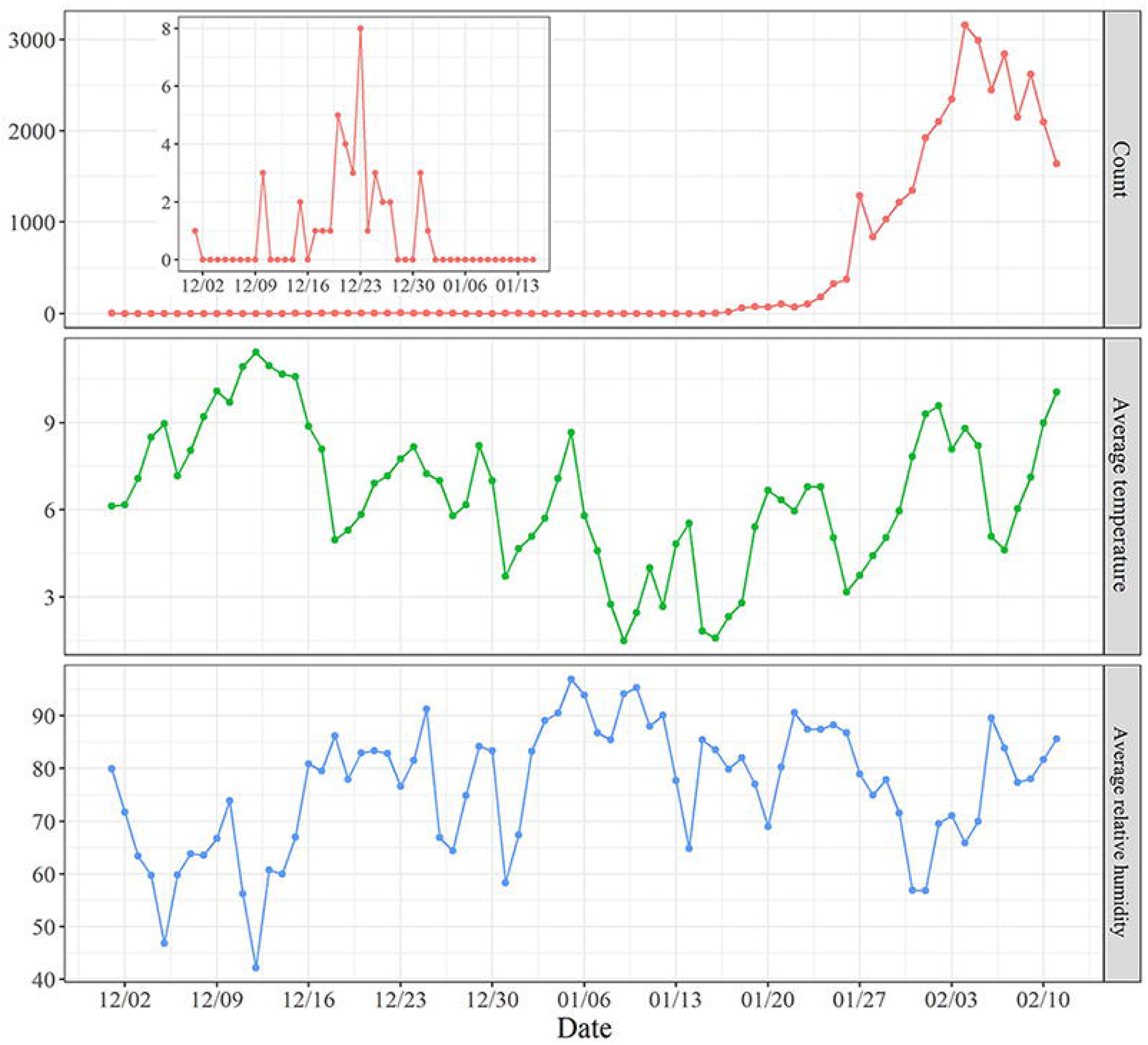
The time series of the daily counts, daily AT, and daily ARH in Hubei province from December 1, 2019 to February 11, 2020.

The estimates of regression coefficients of the GAM for Hubei province are listed in Table 1. There was a significant interaction between AT and ARH (0.04, 95% confidence interval (CI): 0.004–0.07), and the effect of the Baidu index was also statistically significant. Significant interactions between AT and ARH were also found in Zhejiang, Shandong, Hebei, Jilin, and Gansu (Table A2).

The associations of meteorological factors and the daily count of COVID-19 for Hubei province are visualized in Fig. 3. With ARH fixed at its 25^th^, 50^th^, and 75^th^ percentiles, AT showed a negative association with COVID-19, with the IRRs being 0.43 (95% CI: 0.25–0.73), 0.5 (95% CI: 0.32–0.78), and 0.64 (95%CI: 0.43–0.94), respectively. Similarly, there was a negative relationship between ARH and COVID-19 with the AT fixed at its 25^th^, 50^th^, and 75^th^ percentiles. The IRRs here were 0.78 (95% CI: 0.69–0.89), 0.84 (95% CI: 0.75–0.93), and 0.89 (95% CI: 0.8–1). Every 1°C increase in AT led to a decrease in the daily confirmed cases by 36% to 57% when ARH was in 67% to 85.5% and every 1% increase in ARH led to a decrease in the daily confirmed cases by 11% to 22% when AT was 5.04°C to 8.2°C.

**Fig 3.**
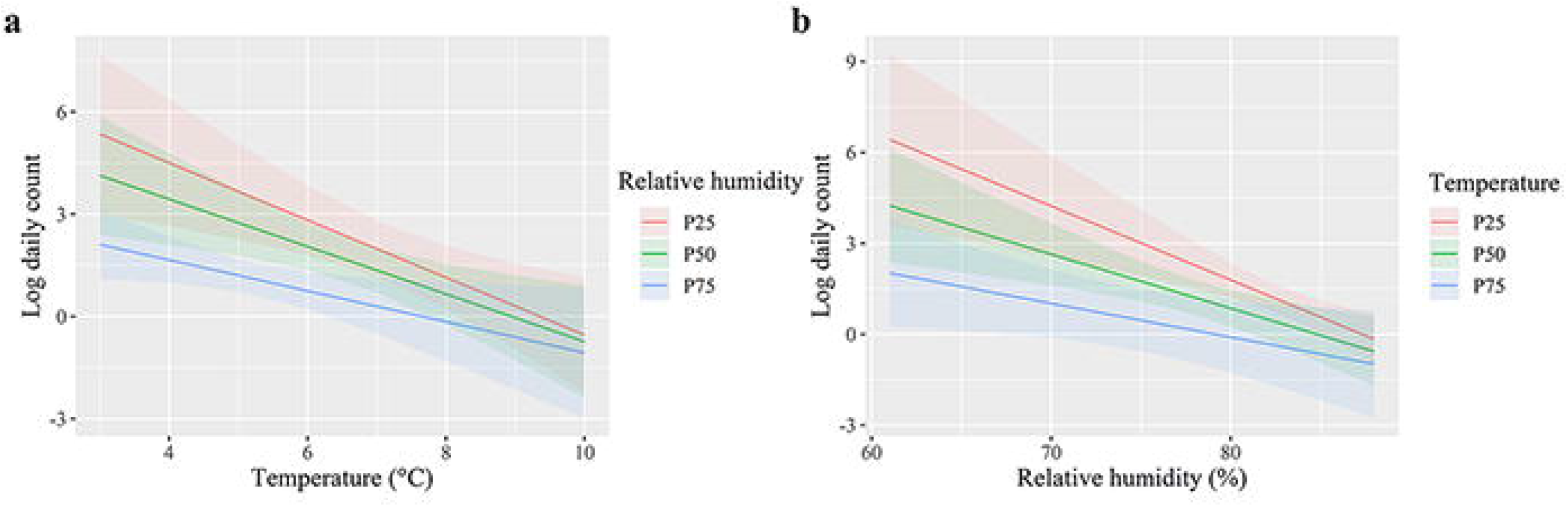
Effect plots for the impact of (a) AT and (b) ARH on the daily counts of COVID-19.

Fig. 4 shows the IRRs for AT in association with COVID-19 when ARH was fixed at its median for each province. There was also a significant positive effect of AT in Hunan, Anhui, Jiangsu, Chongqing, Liaoning, and Jilin in addition to Hubei. However, AT showed an opposite effect in the nearby provinces of Hubei and two provinces in northeast China. We conducted a sensitivity analysis by fixing ARH at its 25^th^ and 75^th^ percentiles (Figs. A4 and A5), and the variation in the association of AT with COVID-19 with ARH alteration was not substantial.

**Fig 4.**
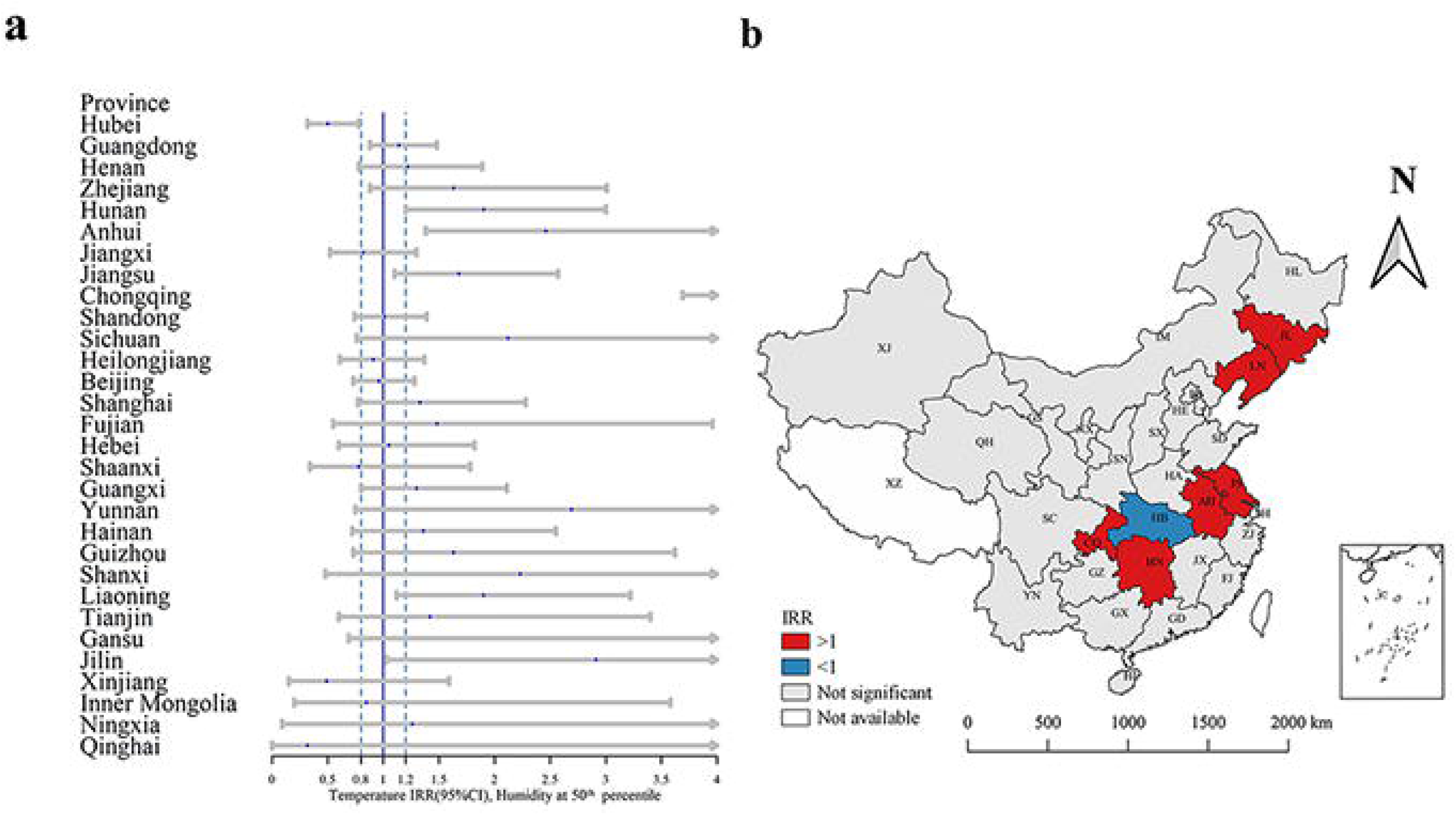
The forest plot (a) and spatial distribution (b) of IRRs of AT with ARH fixed at its median in all the provinces in China.

Fig. 5 shows the IRRs for ARH in association with COVID-19 when AT was fixed at its median for each province. Unlike Hubei province, other provinces including Henan, Anhui, and Chongqing had a positive relationship between ARH and COVID-19. The estimates of IRRs and the forest plots of the ARH with AT fixed at its 25^th^ and 75^th^ percentiles as a sensitivity analysis are presented in Figs. A6 and A7, and there were notable variations in the relationship between ARH and COVID-19 at the different levels of AT.

**Fig 5.**
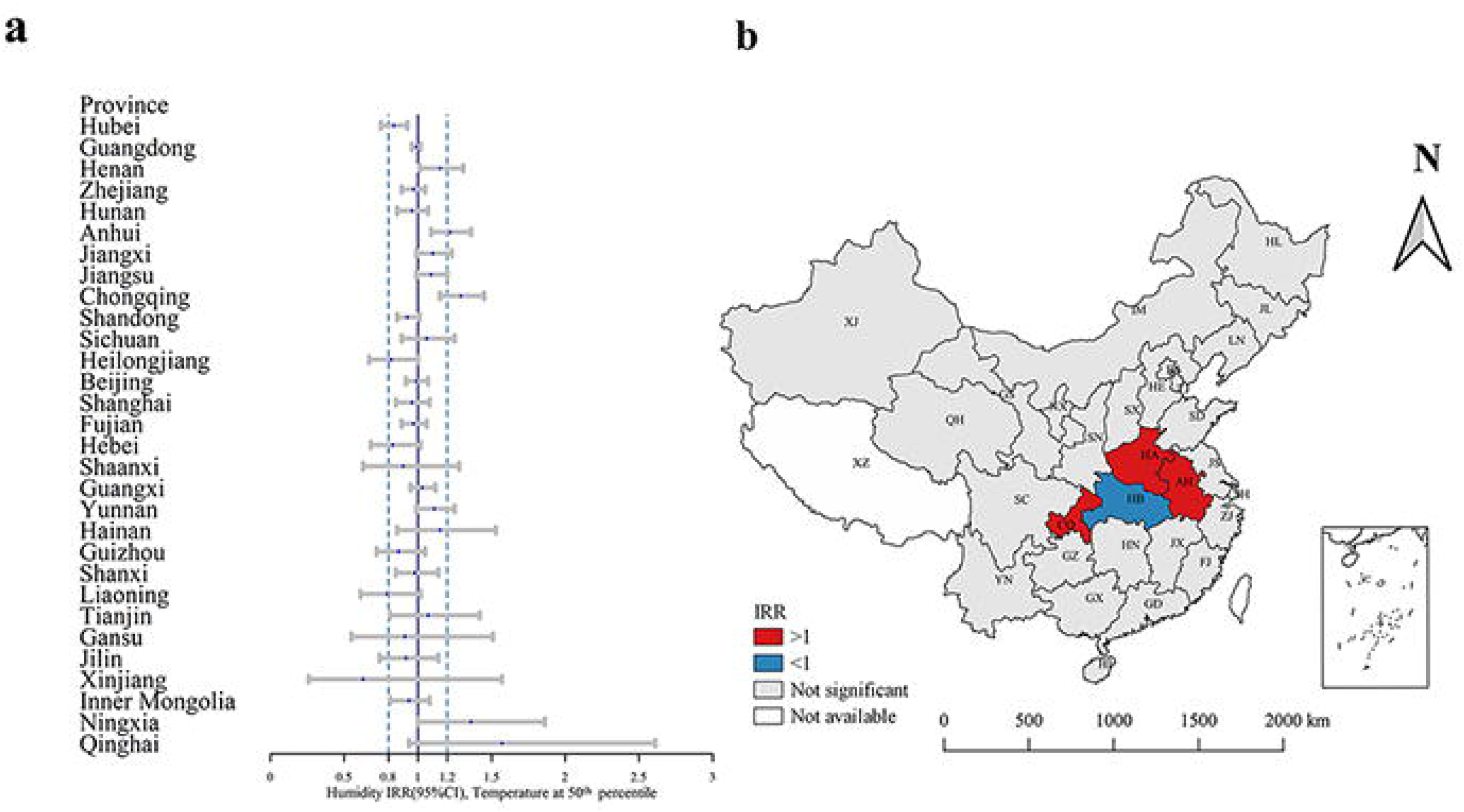
The forest plot (a) and spatial distribution (b) of IRRs of ARH with AT fixed at its median in all provinces in China.

## Discussion

Our study suggests that both daily temperature and relative humidity influenced the occurrence of COVID-19 in Hubei province and in some other provinces. However, the association between COVID-19 and AT and ARH across the provinces was not consistent. We found spatial heterogeneity of COVID-19 incidence, as well as its relationship with daily AT and ARH, among provinces in Mainland China.

The study period − and hence time series length − was longer for Hubei province than other provinces. The longer the study period, the more stable the model results are expected to be. Considering the incubation period of COVID-19, we used the 14-day EMA of daily AT and ARH to investigate the effects of ARH and AT on COVID-19. Most of the confirmed cases (and therefore meteorological data) were derived from the period before travel restrictions were enacted for Wuhan city (January 23, 2020); therefore, the Hubei model most likely reflected the real (unconfounded) effect of meteorological factors on COVID-19, where negative correlations between AT and ARH and the incidence of COVID-19 were found. Evidence from previous epidemiological studies and laboratory investigations have documented negative associations between ARH and AT and other coronavirus-related diseases [9, 14, 15]. One study reported that the risk of an increased daily incidence of SARS was 18.18 (95% CI: 5.6–58.8) times greater at lower temperature during the outbreak than on warmer days[16]. SARS-CoV[17] and MERS-CoV[9, 18] are more stable in cold and dry conditions; low temperature and low humidity lead to an increase of suspended matter in the atmosphere, facilitating ideal conditions for virus attachment, replication and transmission, while low temperature can also dry out the mucous membrane, reduce the function of cilia, and support the survival and transmission of the virus and the spread of diseases[6, 8, 14, 19, 20].

A novel finding of this study is the significant interaction between ARH and AT, and COVID-19 transmission. In Hubei province, the interaction between ARH and AT was found to have a positive effect on transmission (0.04, 95% CI: 0.004–0.07). Increased AT (ARH) led to a decreased effect of ARH (AT) on the incidence of COVID-19 in Hubei province. The exact mechanism of the interaction is unclear. One probable reason might be that a combination of low AT and humidity make the nasal mucosa prone to small ruptures, creating opportunities for virus invasion[21]. A previous study observed an interaction between minimum temperature and humidity on influenza [22, 23]. However, previous studies have seldom analyzed the effects of the interaction of meteorological factors, focusing on the single effect of humidity or temperature on the development of COVID-19[24, 25]. Hence, when estimating the impact of climate on the risk of COVID-19 transmission, the interaction between meteorological factors should be considered. Research findings on meteorological factors and their interactions should be incorporated into the prevention and control of COVID-19. With the arrival of spring in China, the increased AT in the southern region might lead to a decrease in the incidence of the disease. Conversely, in northern regions with continuing low ATs and low ARH levels there should be more attention given to monitoring and prevention of COVID-19 due to continuing suitable climatic conditions in these regions.

The effects of AT and ARH showed spatial heterogeneity and there are several possible reasons. The between-region variability of meteorological factors is likely the main reason for the heterogeneity of effects. Small sample size in some provinces leads to model instability and invalid estimates. Most of the daily count of confirmed cases in provinces other than Hubei included in our models (68.6%)[26] were identified as imported cases from Hubei province or cases having a close contact with imported cases for the study period. Meteorological factors mainly affect COVID-19 transmission by affecting the survival of the coronavirus in the environment, and we assume that the coronavirus was unlikely to exist in the environment in other provinces, where most cases were imported during the study period. The city of Wuhan was forced to shut down both outbound and inbound traffic on January 23, 2020, and other provinces then also gradually placed restrictions on the movement of their populations. These intervention measures are expected to also have an important impact on the associations between meteorological factors and the transmission of the virus. However, because of the interventions that were progressively imposed during January and February 2020, the least biased (confounded) time series of COVID-19 cases is that derived from Wuhan and Hubei province, which showed a negative relationship between AT and ARH and cases, with a positive interaction between AT and ARH.

There are some limitations in our study. First, some potential risk factors that could impact the incidence of COVID-19 − for example, province-specific social-economic status − were not included in the model. The study period is short, so that it can be assumed that many covariates did not vary substantially during such a short time period. Second, the incidence of COVID-19 in provinces other than Hubei was more likely to be influenced by interventions after the outbreak in Hubei province. In addition, the meteorological data were collected from the Weather Underground for the capital city for each province only, and data accuracy and representativeness can be improved in future studies. Provinces (except for Hubei) had a short study period and included many imported cases.

In conclusion, meteorological factors influence COVID-19 transmission and spread, potentially with an interactive effect between daily temperature and relative humidity on COVID-19 incidence. There were spatial and temporal heterogeneities in COVID-19 occurrence, which could be attributed to meteorological factors as well as interventions measures across provinces. The reasons for the inconsistency in the impact of meteorological factors on COVID-19 among provinces needs further study.

## Data Availability

Our data were collected from online or public databases. Daily counts of laboratory-confirmed cases were collected from the official reports of the National Health Commission of People’s Republic of China. The meteorological data were retrieved from Weather Underground. The Baidu index were collected from Baidu (the largest search engine in China).

https://www.wunderground.com/

http://index.baidu.com/

http://en.nhc.gov.cn/

## References

1. Wu JT, Leung K, Leung GM: Nowcasting and forecasting the potential domestic and international spread of the 2019-nCoV outbreak originating in Wuhan, China: a modelling study. Lancet (London, England) 2020.

2. Huang C, Wang Y, Li X, Ren L, Zhao J, Hu Y, Zhang L, Fan G, Xu J, Gu X et al: Clinical features of patients infected with 2019 novel coronavirus in Wuhan, China. Lancet (London, England) 2020, 395(10223):497–506.

3. Situation reports of COVID-19 in China: 02 March, 2020 [http://www.nhc.gov.cn/xcs/yqtb/202003/c588ee20113b4136b27f2a07faa7075b.shtml]

4. WHO characterizes COVID-19 as a pandemic [https://www.who.int/emergencies/diseases/novel-coronavirus-2019/events-as-they-happen]

5. Diagnosis and Treatment Protocol of COVID-19 (the 6th Tentative Version) [http://www.nhc.gov.cn/yzygj/s7653p/202002/8334a8326dd94d329df351d7da8aefc2/files/b218cfeb1bc54639af227f922bf6b817]

6. Casanova LM, Jeon S, Rutala WA, Weber DJ, Sobsey MD: Effects of air temperature and relative humidity on coronavirus survival on surfaces. Appl Environ Microbiol 2010, 76(9):2712–2717.

7. Cai QC, Lu J, Xu QF, Guo Q, Xu DZ, Sun QW, Yang H, Zhao GM, Jiang QW: Influence of meteorological factors and air pollution on the outbreak of severe acute respiratory syndrome. Public Health 2007, 121(4):258–265.

8. Tan JG, Mu LN, Huang JX, Yu SZ, Chen BH, Yin J: An initial investigation of the association between the SARS outbreak and weather: with the view of the environmental temperature and its variation. Journal of Epidemiology and Community Health 2005, 59(3):186–192.

9. Gardner EG, Kelton D, Poljak Z, Van Kerkhove M, von Dobschuetz S, Greer AL: A case-crossover analysis of the impact of weather on primary cases of Middle East respiratory syndrome. BMC Infect Dis 2019, 19(1):113.

10. Altamimi A, Ahmed AE: Climate factors and incidence of Middle East respiratory syndrome coronavirus. J Infect Public Health 2019.

11. Chinese Center for Disease Control and Prevention. Epidemic update and risk assessment of 2019 Novel Coronavirus [http://www.chinacdc.cn/yyrdgz/202001/P020200128523354919292.pdf]

12. Li Q, Guan X, Wu P, Wang X, Zhou L, Tong Y, Ren R, Leung KSM, Lau EHY, Wong JY et al: Early Transmission Dynamics in Wuhan, China, of Novel Coronavirus–Infected Pneumonia. New England Journal of Medicine 2020, 382(13):1199–1207.

13. Du Z, Xu L, Zhang W, Zhang D, Hao Y: Predicting the hand, foot, and mouth disease incidence using search engine query data and climate variables: An ecological study in Guangdong, China. Bmj Open 2017, 7(10):e016263.

14. Chan KH, Peiris JS, Lam SY, Poon LL, Yuen KY, Seto WH: The Effects of Temperature and Relative Humidity on the Viability of the SARS Coronavirus. Adv Virol 2011, 2011:734690.

15. Zhang Qiang YX-w, YE Dian-xiu, XIAO Feng-jin, CHENG Zheng-hong: Meteorological Characteristics and Their Impacts during the SARS Epidemic Period. Journal of Nanjing Institute of Meteorology 2004(06):849–855.

16. Lin K, Fong D, Zhu B, Karlberg J: Environmental factors on the SARS epidemic: Air temperature, passage of time and multiplicative effect of hospital infection. Epidemiology & Infection 2006, 134(2):223–230.

17. Cai QC, Jiang QW, Zhao GM, Guo Q, Cao GW, Chen T: Putative caveolin-binding sites in SARS-CoV proteins. Acta pharmacologica Sinica 2003, 24(10):1051–1059.

18. van Doremalen N, Bushmaker T, Munster VJ: Stability of Middle East respiratory syndrome coronavirus (MERS-CoV) under different environmental conditions. Eurosurveillance 2013, 18(38):7–10.

19. Guionie O, Courtillon C, Allee C, Maurel S, Queguiner M, Eterradossi N: An experimental study of the survival of turkey coronavirus at room temperature and +4 degrees C. Avian Pathol 2013, 42(3):248–252.

20. Lowen A, Palese P: Transmission of influenza virus in temperate zones is predominantly by aerosol, in the tropics by contact: a hypothesis. PLoS Curr 2009, 1:RRN1002.

21. Zhou Z-X, Jiang C-Q: Effect of environment and occupational hygiene factors of hospital infection on SARS outbreak. Zhonghua Lao Dong Wei Sheng Zhi Ye Bing Za Zhi 2004, 22(4):261–263.

22. Liu Z, Zhang J, Zhang Y, Lao J, Liu Y, Wang H, Jiang B: Effects and interaction of meteorological factors on influenza: Based on the surveillance data in Shaoyang, China. Environmental Research 2019, 172:326–332.

23. Firestone SM, Cogger N, Ward MP, Toribio J-ALML, Moloney BJ, Dhand NK: The Influence of Meteorology on the Spread of Influenza: Survival Analysis of an Equine Influenza (A/H3N8) Outbreak. PLOS ONE 2012.

24. Luo W, Majumder MS, Liu D, Poirier C, Mandl KD, Lipsitch M, Santillana M: The role of absolute humidity on transmission rates of the COVID-19 outbreak. medRxiv 2020:2020.2002.2012.20022467.

25. Wang M, Jiang A, Gong L, Luo L, Guo W, Li C, Zheng J, Li C, Yang B, Zeng J et al: Temperature significant change COVID-19 Transmission in 429 cities. medRxiv 2020:2020.2002.2022.20025791.

26. The Novel Coronavirus Pneumonia Emergency Response Epidemiology T: The Epidemiological Characteristics of an Outbreak of 2019 Novel Coronavirus Diseases (COVID-19) — China, 2020. China CDC Weekly 2020, 2(8):113–122.

